# Examining the epigenetic mechanisms of childhood adversity and sensitive periods: a gene set-based approach

**DOI:** 10.1101/2021.06.22.21259356

**Authors:** Yiwen Zhu, Alexandre A. Lussier, Andrew D.A.C. Smith, Andrew J. Simpkin, Matthew J. Suderman, Esther Walton, Erin C. Dunn

## Abstract

**Background:** Sensitive periods are developmental stages of heightened plasticity when exposure to childhood adversity may exert lasting impacts. A few biological pathways are known to play key roles in regulating sensitive period plasticity across brain development. Epigenetic mechanisms including DNA methylation (DNAm) may provide a means through which life experiences during sensitive periods induce long-term biological changes. In the current study, we investigated the possibility that adversity during sensitive periods led to DNAm changes in genes that regulate the timing and duration of sensitive periods in development.

**Methods:** Using childhood adversity data and genome-wide DNAm profiles from the Avon Longitudinal Study of Parents and Children (n=785), we summarized DNAm variation of CpG sites in the promoters of genes regulating sensitive periods with the first two principal components (PCs). DNAm summaries were calculated for genes regulating sensitive period opening (n_genes_=15), closing (n_genes_=36), and expression/duration (n_genes_=8). We then performed linear discriminant analysis to test associations between these DNAm summaries and the timing of exposure to seven types of adversity.

**Results:** Sexual or physical abuse and financial hardship during middle childhood (6-7 years) were associated with DNAm of genes regulating the onset and duration of sensitive periods. Sensitivity analyses assessing the presence of any exposure before age 7 and a composite measure of adversity yielded fewer signals, highlighting the importance of accounting for timing and adversity type.

**Conclusions:** With our novel gene set-based approach, we have uncovered suggestive evidence that epigenetic regulation of developmental plasticity may be affected by early life adversity. The complementarity of our gene-level view of the epigenome to the more common and granular epigenome-wide association study may yield novel mechanistic insights not only for adversity but also for other exposures and outcomes.

## 1. Introduction

Early life exposure to adversity can exert profound impacts on development, leading to changes in myriad neurological, behavioral, and health outcomes (Berens et al., 2017; Oh et al., 2018). Animal and human studies have suggested these adverse effects may be time-varying, with some developmental stages being sensitive periods of increased plasticity and vulnerability to the effects of adversity (Nelson and Gabard-Durnam, 2020).

Genetic pathways play a key role in regulating the timing of sensitive periods. Studies in animals and humans have identified more than 50 genes involved in shaping the *opening, closing*, and *expression* or *duration* of sensitive period plasticity across brain development (Mauney et al., 2013; Takesian and Hensch, 2013). These mechanisms have been identified in both cortical regions and subcortical structures (e.g., amygdala and hippocampus) (Gogolla et al., 2009).

Emerging evidence suggests that childhood adversity during certain developmental periods may have long-term effects on DNA methylation (DNAm) at multiple loci across the genome (Dunn et al., 2019). Given the well-known relationship between DNAm and gene expression, these changes could mediate the lasting impacts of childhood adversity. Research in developmental neuroscience supports the hypothesis that trajectories of sensitive periods and brain development may be sensitive to epigenetic changes including changes in DNAm (Reh et al., 2020). However, evidence for whether and how the timing of adversity shapes epigenetic modifications of pathways regulating sensitive periods in human populations is lacking.

Examining epigenetic mechanisms of childhood adversity and sensitive periods may provide new insights into functional changes underlying the pathophysiology of neuropsychiatric disorders (Do et al., 2015).

To address these gaps, we analyzed data from a unique population-based sample to identify if and when childhood adversity exerted strong effects during the first seven years of life on DNAm of three sensitive period gene pathways. The gene sets were identified from a literature review of existing animal and human studies on known genetic components implicated in the regulation of the *opening* (n_genes_=15), *closing* (n_genes_=36), and *expression* (n_genes_=8) of sensitive period functioning across brain development (**Figure 1**; Takesian and Hensch, 2013; Zhu et al., 2021). We used a novel analytic approach to assess the relationship between time-varying adversity and summaries of DNAm variation at the gene set level.

**Figure 1.**
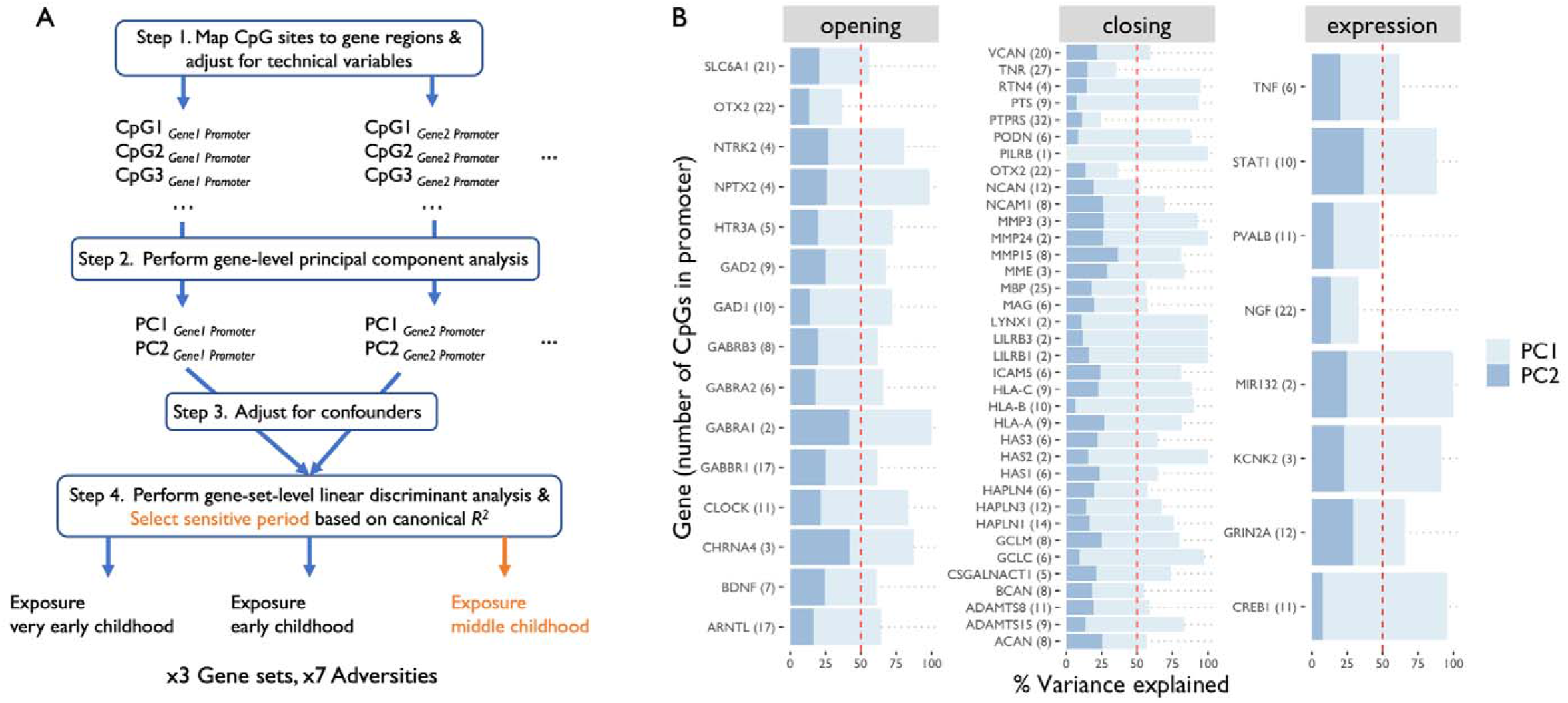
**A) Flowchart of analyses.** After adjusting for technical variables (step 1), variation among CpG sites annotated to the promoter of a given gene was summarized at the *gene* level into two principal component (PC) scores (step 2). PCs were then adjusted for confounders (step 3). Finally, at the *gene set* level, associations between time-varying exposures to childhood adversity and PCs of all genes in a given gene set were assessed using a linear discriminant analysis (step 4). The sensitive period most strongly associated with the PCs was selected based on canonical as highlighted in orange in this example. **B) Variances of CpGs located in promoters of three sets of sensitive period genes, as explained by the first two principal components (PCs)**. The number of CpGs annotated to the promoter of each gene is noted in parentheses. The red dashed line indicates 50% of the variance. For 53 of 58 genes, variance explained by the first two PCs exceeded 50%.

**Table 1.**
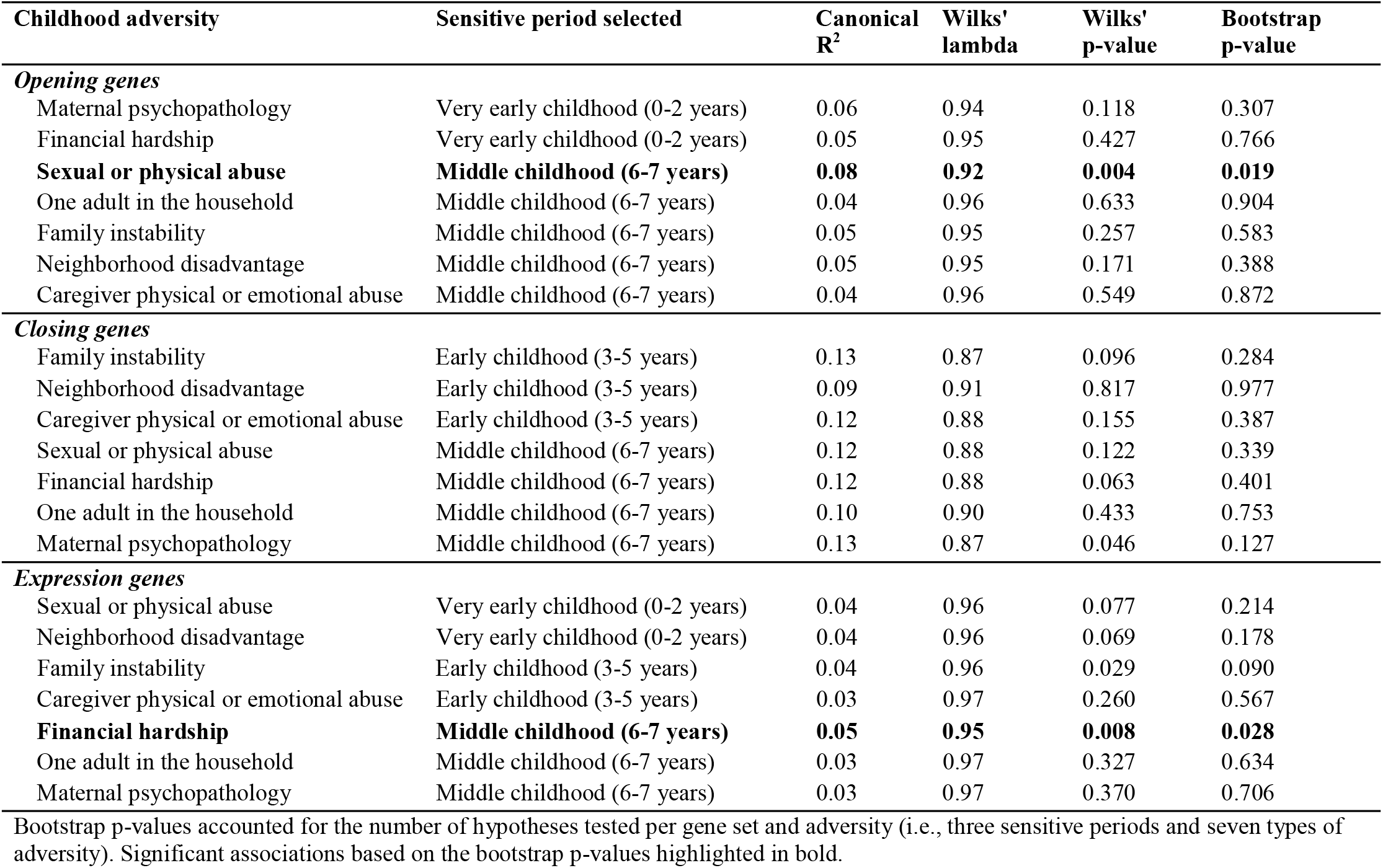
Associations between time-dependent exposure to childhood adversity and DNA methylation at promoters of three gene sets regulating sensitive period functioning.

## 2. Materials and Methods

### 2.1 Sample and Procedures

Data came from the Accessible Resource for Integrated Epigenomics Studies (ARIES) (Relton et al., 2015), a subsample of a UK-based prospective birth cohort, the Avon Longitudinal Study of Parents and Children (ALSPAC) (Boyd et al., 2013; Fraser et al., 2013). ARIES consists of 1,018 mother-child pairs with data across five waves of data collection, including blood-based, genome-wide DNAm profiles. Our analytic sample included 785 singleton child participants who had DNAm data at age 7, as well as complete data on all covariates and at least one type of adversity across all time points (**Supplemental Materials**).

### 2.2 Measures

#### 2.2.1 Exposure to Childhood Adversity

We analyzed seven common types of childhood adversity, described in detail elsewhere (Dunn et al., 2019) and summarized in **Table S1**: (a) caregiver physical or emotional abuse; (b) sexual or physical abuse (by anyone); (c) maternal psychopathology; (d) one adult in the household; (e) family instability; (f) financial hardship; and (g) neighborhood disadvantage.

These measures were captured from maternal reports at four or more time points between birth and age 7.

To assess sensitive period effects of adversity on DNAm, we grouped children based on the presence or absence of exposure during each of three developmental periods: very early childhood (0-2 years), early childhood (3-5 years), and middle childhood (6-7 years).

#### 2.2.2 DNA Methylation

DNAm profiles were obtained from white blood cell samples collected around age 7 (Relton et al., 2015). DNAm was assayed using the Illumina Infinium Human Methylation 450k BeadChip microarray. Beta values in DNAm were analyzed, which represented the proportion of cells methylated at each probe. Functional normalization was used to remove noise due to technical variations (Min et al., 2018). Prior to analysis, duplicate samples, polymorphic probes, and XY-binding cross-hybridizing probes were removed. Winsorization was performed to reduce the impact of DNAm outliers.

We focused on DNAm in promoter regions, which is known to have more robust associations with transcription levels (Vialou et al., 2013). Promoters were defined as the genomic regions upstream of the transcription start sites of genes (TSS1500, TSS200) and from 5’-untranslated region through the first exon (Lokk et al., 2014). A total of 530 CpG sites were annotated to the promoters of the 58 sensitive period regulating genes. The number of CpG sites annotated to each gene promoter varied greatly, ranging from n=1 (*PILRB*) to n=31 (*PTPRS*) (**Figure 1B**).

#### 2.2.3 Covariates

To control for potential confounding by sociodemographic characteristics that could influence both exposure to childhood adversity and DNAm, we adjusted for the following covariates: child race/ethnicity (White; non-White); child birth weight (continuous, grams); maternal age (ages 15-19; ages 20-35; ages 35+); number of previous pregnancies (0;1;2; 3+); sustained maternal smoking during pregnancy (yes; no); and maternal education at baseline (levels corresponding to below age 16, age 16, post age 16, and post age 18 qualifications).

### 2.3 Data Analysis

The analytic steps are shown in **Figure 1A**. First, using principal components analysis (PCA), we summarized promoter DNAm variation for each gene. Prior to PCA, technical variables were regressed from DNAm levels at each CpG site (estimated cell proportions and sample type). Two first two principal components were retained for each gene.

Associations between these principal components and adversity were tested using linear discriminant analysis (LDA). LDA computes a linear combination of features, in this case DNAm principal components for a gene, that maximally separates two groups, in this case exposed versus unexposed individuals. The statistical strength of this separation is calculated as the squared canonical correlation (canonical R^2^), which is the multivariate equivalent of a Pearson’s correlation coefficient. Among the exposures encoding sensitive periods of very early childhood, early childhood, and middle childhood, the exposure with the largest canonical R^2^ was selected and a p-value was calculated through a bootstrap procedure with 5000 iterations, which accounted for testing multiple hypotheses.

Finally, we performed sensitivity analyses to test whether genomic regions, adversity timing, or adversity type mattered. We also compared the gene set-based findings of the current study to results obtained from a previous genome-wide CpG-level analysis examining time-varying exposures to adversity in ALSPAC (Lussier et al., 2021). Details of the sensitivity analyses are summarized in **Supplemental Materials**.

## 3. Results

### 3.1 Sample Characteristics

Compared to the entire ALSPAC sample, children in the analytic sample (n=785) were more likely to be born to mothers who were older, had fewer previous pregnancies, less likely to smoke during pregnancy, and better educated at baseline. Children in the analytic sample were also less likely than the larger cohort to experience childhood adversity (**Table S2**). The prevalence of childhood adversity ranged from 2.2% to 24% across adversities and time periods (**Figure S1**). Exposures were moderately to strongly correlated across time periods within a adversity (**Figure S2**), and weakly to moderately correlated across adversities within a period (**Figure S3**).

### 3.2 Principal Component Analysis, Linear Discriminant Analysis, and Model Selection

For 53 of 58 genes, the first two PCs accounted for more than 50% of the variation in promoter DNAm (**Figure 1B**).

Childhood adversity during middle childhood (6-7 years) was most frequently associated with DNAm in the promoters of genes involved in regulating sensitive period functioning domains (selected for 12 out of the 21 LDA models). After bootstrapping to account for having selected the sensitive period with the largest canonical R^2^ for each gene set and adversity, we identified two associations between the timing of childhood adversity and DNAm of relevant gene sets at a bootstrap p-value*<0*.*05*. Specifically, exposure to sexual or physical abuse in middle childhood was associated with significant differences in DNAm among *opening* genes, meaning genes regulating the onset of sensitive periods (p=0.019). Exposure to financial hardship in middle childhood was also associated with *expression* genes, meaning genes regulating the duration of sensitive periods (p=0.028).

Results from sensitivity analyses offered three additional insights. First, DNAm signals were confined to promoter regions only, as no association was identified in the analyses of CpGs annotated to gene bodies (**Table S3**). Second, when lifetime exposure before age 7 was assessed (i.e., ever vs. never exposed), only sexual or physical abuse was associated with DNAm of the *opening* gene promoters (**Table S4**). Third, no significant associations emerged in analyzing a composite measure of childhood adversity before age 7, indicating that the relationships were adversity-type specific (**Table S5**).

Additionally, the CpG-level lookup of results shown by Lussier et al. did not yield much overlap with the current gene set-based analyses (2021). We identified no significant association at FDR<0.05 among the 530 CpGs annotated to promoters of genes regulating sensitive period functioning (**Table S6**), suggesting that the gene set-based method uncovered more signal than CpG-level assessments.

## 4. Discussion

In the current study, we implemented a novel analytic approach to interrogate the joint contribution of time-varying exposures to childhood adversity and epigenetic modifications of pathways implicated in sensitive period functioning. Two significant associations emerged, both of which suggest that exposure to adversity during middle childhood (ages 6-7), namely physical or sexual abuse and financial hardship, is linked to different profiles of childhood DNAm in the promoters of genes involved in the onset or duration of sensitive periods. Middle childhood is a critical stage of development: key structures of neural plasticity, such as perineuronal nets, reach maturation around age 8 in brain regions implicated in psychopathology, including the medial prefrontal cortex and hippocampus (Mauney et al., 2013; Rogers et al., 2018). Adverse environmental stimuli during this period of rapid growth and maturation could lead to disruptions of experience-expectant mechanisms. Epigenetic modifications might occur to downregulate the activity of genes involved in neuronal plasticity, preventing further heightened molecular and behavioral responses. Replication of these findings in larger, more diverse samples, as well as studies focusing on cascading effects later in life, are needed to further disentangle the epigenetic mechanisms of childhood adversity and sensitive periods.

Consistent with prior epigenome-wide studies (Dunn et al., 2019), our study sheds light on the benefits of adopting more nuanced characterization of exposures in two aspects. First, the sensitive period effects of childhood adversity on DNAm confirm that the time-sensitive nature of plasticity must be addressed in developmental research, as opposed to a traditional classification of the absence versus presence of exposure. Second, we also found evidence for differential effects of specific types of childhood adversity. Early life stress in different forms may produce unique biological signatures and alter functioning differently even within the domain of sensitive period regulation. As demonstrated here, examining biological signatures such as DNAm differences of specific pathways may inform the discussion on “lumping vs. splitting” in childhood adversity conceptualization (Smith and Pollak, 2021).

This study has several limitations. First, the gene sets we included were not exhaustive of all possible sensitive period pathways. As the field grows to identify more molecular components regulating sensitive periods, our analyses can be expanded to include additional genes or pathways that relate to specific brain regions. Moreover, ALSPAC participants were predominantly of European ancestry and the subsample with epigenetic data had lower levels of adversity compared to the entire sample. It remains an important objective of future work to examine the generalizability of the current findings in children from minority groups, who often disproportionally experience childhood adversity (Slopen et al., 2016).

In conclusion, our study provides insights into the role of epigenetic mechanisms underlying neural plasticity in the interplay between biological programming and early life adversity. Leveraging the applicability of functionally grouped gene sets, this analytic approach offers a useful complement to current epigenome-wide analyses and could ultimately improve the interpretability of molecular findings in neuropsychiatric research.

## Data Availability

Data are available to approved investigators through the ALSPAC. Please note that the study website contains details of all the data that is available through a fully searchable data dictionary and variable search tool (http://www.bristol.ac.uk/alspac/researchers/our-data/).

## Acknowledgments

We are extremely grateful to all the families who took part in this study, the midwives for their help in recruiting them, and the whole ALSPAC team, which includes interviewers, computer and laboratory technicians, clerical workers, research scientists, volunteers, managers, receptionists and nurses.

