## Supplementary Materials for "Examining the epigenetic mechanisms of childhood adversity and sensitive periods: a gene set-based approach"

**Supplemental Materials**

**Supplementary Methods**

**1. ALSPAC Sample and Data Collection**

The Avon Longitudinal Study of Parents and Children (ALSPAC) is a longitudinal population-based cohort based in Avon, UK. Pregnant individuals residing in the area with estimated delivery dates between April 1, 1991, and December 31, 1992, were invited to participate in the study. The initial sample included 14,514 women who returned at least one questionnaire or attended one focus clinic by July 19, 1999. Out of the initial sample, a total of 14,676 fetuses and 13,988 children who were alive at 1 year of age were included in the study. A second wave of participants were recruited when the oldest children in the cohort reached approximately 7 years of age, which consisted of eligible individuals who did not join the study initially, resulting in the enrollment of an additional 913 children. With the two waves combined, the ALSPAC sample consisted of 14,901 children alive at age 1 year (Boyd et al., 2013; Fraser et al., 2013).

The analytic sample came from a subsample of ALSPAC, the Accessible Resource for Integrated Epigenomics Studies (ARIES). The subsample consisted of 1,018 mother-child pairs from whom blood-based DNA methylation data were collected. Participants in the ARIES subsample were randomly selected from ALSPAC participants with complete data across at least five timepoints of data collection (Relton et al., 2015).

Ethical approval for the study was obtained from the ALSPAC Ethics and Law Committee and the Local Research Ethics Committee. Consent for biological samples has been collected in accordance with the Human Tissue Act (Human Tissue Act, 2004). Please note that the study website contains details of all the data that is available through a fully searchable data dictionary and variable search tool (http://www.bristol.ac.uk/alspac/researchers/our-data/).

**2. DNA Methylation**

DNAm data analyzed in this study were measured at 485,577 CpG sites using the Illumina Infinium Human Methylation 450K BeadChip microarray (Illumina, San Diego, CA). The 450K microarray captures DNAm variation at 99% of RefSeq genes. To minimize batch effects, samples in ARIES were semi-randomly assigned across time points to different slides (Relton et al., 2015). Data quality control was performed based on protocol described by Min et al. (Min et al., 2018). The pipeline applied functional normalization and background correction using the *Meffil* R package. Additionally, cross-hybridizing probes, polymorphic probes, and probes located in sex chromosomes were removed before analyses. To reduce the impact of outliers, beta values at each CpG site were winsorized by setting values outside the 5^th^ and 95^th^ percentiles to the 5^th^ and 95^th^ cutoffs, respectively.

**3. Sensitivity analysis**

To examine whether alternative analytic approaches would lead to the same findings, we pursued the following four sets of sensitivity analyses:

First, we examined whether genomic regions considered in the analyses would impact the results. Specifically, we tested whether time-varying exposures to the same seven types of childhood adversity were associated with DNAm variations in loci located in the gene bodies of the three sets of sensitive period-regulating genes, as opposed to the promoters. The same analytic procedure was followed: beta values at CpG sites in each gene body were summarized into two principal component scores per gene, and a linear discriminant analysis model was fit for each exposure time point. Results are shown in **Table S3**.

Second, we examined whether the consideration of exposure timing mattered in the analyses. Here, we assessed whether the presence of exposure to each type of adversity before age 7, compared to the absence of the given type of exposure, was associated with DNAm variations in promoters of sensitive period gene sets. Again, we pursued the same analytic approach, except that no selection of time period was needed, since only one exposure variable needed to be considered for each adversity type. Results are shown in **Table S4**.

Third, we investigated whether aggregating exposure status across the seven types of adversity might change the results. During each developmental period (very early childhood, early childhood, and middle childhood), children exposed to any of the seven types of childhood adversity were coded as exposed, resulting in three models for each gene set (i.e., one for each sensitive period). Results are shown in **Table S5**.

Lastly, we compared CpG -level results of a similar analysis to the gene set-level results of the current study. Specifically, Lussier et al. performed a structured life course modeling approach to compare multiple encoded exposures of childhood adversity and identify the hypothesis most strongly associated with DNAm at each locus across the epigenome using the ALSPAC cohort (Lussier et al., 2021). After correcting for the multiple testing burden associated with testing the 530 CpG sites analyzed in the current study, we extracted results at four CpG sites at a liberal threshold of FDR <0.2, given the lack of CpG-level findings among loci included in the current study. Results are shown in **Table S6**.

Zhu, Y., Simpkin, A.J., Suderman, M.J., Lussier, A.A., Walton, E., Dunn, E.C., Smith, A.D.A.C., n.d. A Structured Approach to Evaluating Life Course Hypotheses: Moving Beyond Analyses of Exposed Versus Unexposed in the Omics Context. Am J Epidemiol. https://doi.org/10.1093/aje/kwaa246

**Supplementary Tables**

| Table S1. Summary of measures of exposure to childhood adversity analyzed in the current study. | | | | |
| --- | --- | --- | --- | --- |
| **Adversity** | **Respondent** | **Instrument or questionnaire items** | **Exposure definition** | **Time points of assessment** |
| **Caregiver physical or emotional abuse** | Mother and partner | 1) your partner was physically cruel to your children; 2) you were physically cruel to your children; 3) your partner was emotionally cruel to your children; 4) you were emotionally cruel to your children | Children were coded as exposed if either the mother, the partner, or both, endorsed any of the items. | 8 months, 1.75 years, 2.75 years, 4 years, 5 years, and 6 years |
| **Sexual or physical abuse** | Mother | An item asking whether or not the child had been exposed to either sexual or physical abuse from anyone | Children were coded as exposed if an affirmative response was provided to the item. | 1.5 years, 2.5 years, 3.5 years, 4.75 years, 5.75 years, and 6.75 years |
| **Maternal psychopathology** | Mother | 1) the Crown-Crisp Experiential Index (CCEI), assessing anxiety and depression; 2) the Edinburgh Postnatal Depression Scale (EPDS); and 3) a question asking about suicide attempts in the past 1.5 years | Consistent with prior studies and established cutoffs, children were coded as exposed if one or more of the following criteria was met: 1) CCEI depression score > 9; 2) CCEI anxiety score > 10; 3) EPDS score > 12; or the 4) a suicide attempt since the time of the last interview | 8 months, 1.75 years, 2.75 years, 5 years, and 6 years of age |
| **One adult in the household** | Mother | An item asking about the number of adults (>18 years of age) living in the household | Children were coded as exposed if fewer than two adults were residing in the household. | 8 months, 1.75 years, 2.75 years, 4 years, and 7 years |
| **Family instability** | Mother | Child had 1) been taken into care; 2) been separated from their mother for two or more weeks; 3) been separated from their father for two or more weeks; or 4) acquired a new parent. | Children were coded as exposed if at least two of these events occurred at a single time point. | 1.5 years, 2.5 years, 3.5 years, 4.75 years, 5.75 years, and 6.75 years |
| **Financial hardship** | Mother | The family had difficulty affording the following: 1) items for the child; 2) rent or mortgage; 3) heating; 4) clothing; 5) food. Each of the 5 items was coded on a Likert-type scale (1=not difficult; 2=slightly difficult; 3=fairly difficult; 4=very difficult) | Children were coded as exposed if their mothers reported at least fair difficulty for three or more items at each time point. | 8 months, 1.75 years, 2.75 years, 5 years, and 7 years |
| **Neighborhood disadvantage** | Mother | There were problems in the neighborhood: 1) noise from other homes; 2) noise from the street; 3) garbage on the street; 4) dog dirt; 5) vandalism; 6) worry about burglary; 7) mugging; and 8) disturbance from youth. Response options to each item were: 2=serious problem, 1=minor problem, 0=not a problem or no opinion | A sum score was derived, ranging from 0-16. Children with scores >=8, generally corresponding to the 95th percentile, were classified as exposed. | 1.75 years, 2.75 years, 5 years, and 7 years |

| Table S2. Distribution of covariates and prevalence of exposure to childhood adversity in the  analytic sample (n=785), compared to the full ALSPAC sample (n=15646). | | | |
| --- | --- | --- | --- |
|  | ALSPAC full sample | Analytic sample | Comparison |
|  | (n=14901) | (n=785) | p-value |
| ***Covariates*** |  |  |  |
|  | *n (%)* | *n (%)* |  |
| Race |  |  |  |
| Non-White | 611 (5.1) | 24 (3.1) | 0.011 |
| White | 11488 (94.9) | 761 (96.9) |  |
| Sex |  |  |  |
| Male | 7542 (51.3) | 405 (51.6) | 0.908 |
| Female | 7152 (48.7) | 380 (48.4) |  |
| Age of mother at child's birth |  |  |  |
| Ages 15-19 | 650 (4.6) | 4 (0.5) | <0.001 |
| Ages 20-35 | 12363 (88.4) | 698 (88.9) |  |
| Age 36+ | 968 (6.9) | 83 (10.6) |  |
| Number of previous pregnancies |  |  |  |
| 0 | 5800 (44.7) | 357 (45.5) | 0.024 |
| 1 | 4550 (35.0) | 295 (37.6) |  |
| 2 | 1860 (14.3) | 104 (13.2) |  |
| 3+ | 772 (5.9) | 29 (3.7) |  |
| Sustained smoking during pregnancy | |  |  |
| No | 9565 (78.8) | 704 (89.7) | <0.001 |
| Yes | 2577 (21.2) | 81 (10.3) |  |
| Maternal education at baseline |  |  |  |
| Below O-level | 3735 (30.0) | 101 (12.9) | <0.001 |
| O-level | 4303 (34.6) | 268 (34.1) |  |
| A-level | 2795 (22.5) | 244 (31.1) |  |
| Degree or above | 1603 (12.9) | 172 (21.9) |  |
|  | *Mean (SD)* | *Mean (SD)* |  |
| Birth weight, grams | 3391.65 (560.04) | 3488.56 (488.04) | <0.001 |
| ***Exposure prevalence*** |  |  |  |
|  | *n (%)* | n (%) |  |
| Physical or sexual abuse | 1564 (23.4) | 122 (18.2) | 0.001 |
| Financial hardship | 3273 (41.0) | 164 (24.1) | <0.001 |
| One adult in the household | 2336 (32.1) | 108 (16.0) | <0.001 |
| Family instability | 1760 (23.8) | 125 (18.0) | <0.001 |
| Neighborhood disadvantage | 1815 (24.2) | 112 (16.0) | <0.001 |
| Caregiver physical or emotional abuse | 1944 (28.6) | 147 (21.0) | <0.001 |
| Maternal psychopathology | 5350 (60.3) | 297 (43.4) | <0.001 |
| *Note.* P-values were obtained from χ^2^ tests or independent t-tests comparing distributions of covariates and exposure variables in the analytic sample to distributions in the entire ALSPAC sample. | | | |

| Table S3. Associations between time-dependent exposures to childhood adversity and DNA methylation at gene bodies of three gene sets regulating sensitive period functioning. | | | | | |
| --- | --- | --- | --- | --- | --- |
| **Childhood adversity** | **Sensitive period selected** | **Canonical R^2^** | **Wilks' lambda** | **Wilks' p-value** | **Bootstrap**  **p-value** |
| ***Opening genes*** | | | | | |
| Family instability | Very early childhood (0-2 years) | 0.04 | 0.96 | 0.206 | 0.508 |
| Caregiver physical or emotional abuse | Early childhood (3-5 years) | 0.05 | 0.95 | 0.045 | 0.127 |
| Maternal psychopathology | Early childhood (3-5 years) | 0.04 | 0.96 | 0.141 | 0.340 |
| Sexual or physical abuse | Middle childhood (6-7 years) | 0.05 | 0.95 | 0.075 | 0.216 |
| Financial hardship | Middle childhood (6-7 years) | 0.04 | 0.96 | 0.293 | 0.608 |
| One adult in the household | Middle childhood (6-7 years) | 0.03 | 0.97 | 0.735 | 0.960 |
| Neighborhood disadvantage | Middle childhood (6-7 years) | 0.05 | 0.95 | 0.105 | 0.257 |
| ***Closing genes*** | | | | | |
| Sexual or physical abuse | Very early childhood (0-2 years) | 0.10 | 0.90 | 0.211 | 0.522 |
| Neighborhood disadvantage | Very early childhood (0-2 years) | 0.09 | 0.91 | 0.456 | 0.781 |
| Caregiver physical or emotional abuse | Early childhood (3-5 years) | 0.10 | 0.90 | 0.303 | 0.629 |
| Financial hardship | Middle childhood (6-7 years) | 0.11 | 0.89 | 0.190 | 0.449 |
| One adult in the household | Middle childhood (6-7 years) | 0.10 | 0.90 | 0.345 | 0.658 |
| Family instability | Middle childhood (6-7 years) | 0.11 | 0.89 | 0.164 | 0.430 |
| Maternal psychopathology | Middle childhood (6-7 years) | 0.11 | 0.89 | 0.143 | 0.347 |
| ***Expression genes*** |  |  |  |  |  |
| One adult in the household | Very early childhood (0-2 years) | 0.03 | 0.97 | 0.277 | 0.559 |
| Neighborhood disadvantage | Very early childhood (0-2 years) | 0.03 | 0.97 | 0.312 | 0.625 |
| Maternal psychopathology | Early childhood (3-5 years) | 0.03 | 0.97 | 0.249 | 0.535 |
| Sexual or physical abuse | Middle childhood (6-7 years) | 0.03 | 0.97 | 0.127 | 0.331 |
| Financial hardship | Middle childhood (6-7 years) | 0.03 | 0.97 | 0.397 | 0.747 |
| Family instability | Middle childhood (6-7 years) | 0.03 | 0.97 | 0.104 | 0.286 |
| Caregiver physical or emotional abuse | Middle childhood (6-7 years) | 0.04 | 0.96 | 0.032 | 0.098 |
| Bootstrap p-values accounted for the number of hypotheses tested per gene set and adversity (i.e., three sensitive periods). No association was significant. | | | | | |

| Table S4. Associations between being ever exposed to each type of childhood adversity before age 7 and DNA methylation of CpGs annotated to promoters of gene sets regulating sensitive period functioning. | | | |
| --- | --- | --- | --- |
| **Childhood adversity** | **Canonical R^2^** | **Wilks' lambda** | **Wilks' p-value** |
| ***Opening genes*** | | | |
| **Sexual or physical abuse** | **0.06** | **0.94** | **0.041** |
| Financial hardship | 0.04 | 0.96 | 0.658 |
| One adult in the household | 0.03 | 0.97 | 0.882 |
| Family instability | 0.03 | 0.97 | 0.881 |
| Neighborhood disadvantage | 0.04 | 0.96 | 0.532 |
| Caregiver physical or emotional abuse | 0.03 | 0.97 | 0.755 |
| Maternal psychopathology | 0.05 | 0.95 | 0.286 |
| ***Closing genes*** | | | |
| Sexual or physical abuse | 0.11 | 0.89 | 0.212 |
| Financial hardship | 0.08 | 0.92 | 0.867 |
| One adult in the household | 0.09 | 0.91 | 0.549 |
| Family instability | 0.11 | 0.89 | 0.209 |
| Neighborhood disadvantage | 0.07 | 0.93 | 0.879 |
| Caregiver physical or emotional abuse | 0.08 | 0.92 | 0.771 |
| Maternal psychopathology | 0.10 | 0.90 | 0.202 |
| ***Expression genes*** |  |  |  |
| Sexual or physical abuse | 0.01 | 0.99 | 0.851 |
| Financial hardship | 0.03 | 0.97 | 0.243 |
| One adult in the household | 0.02 | 0.98 | 0.707 |
| Family instability | 0.03 | 0.97 | 0.258 |
| Neighborhood disadvantage | 0.02 | 0.98 | 0.554 |
| Caregiver physical or emotional abuse | 0.01 | 0.99 | 0.886 |
| Maternal psychopathology | 0.02 | 0.98 | 0.784 |
| Because only one hypothesis was tested for each type of adversity, no bootstrapping was performed.  Bolded value indicated nominally significant association (p<0.05). | | | |

| Table S5. Associations between being ever exposed to any of the seven types of adversity during each time period and DNA methylation of CpGs annotated to promoters of gene sets regulating sensitive period functioning. | | | | |
| --- | --- | --- | --- | --- |
| **Gene set** | **Sensitive period selected** | **Canonical R^2^** | **Wilks' lambda** | **Wilks' p-value** |
| Opening | Middle childhood (6-7 years) | 0.06 | 0.94 | 0.229 |
| Closing | Middle childhood (6-7 years) | 0.12 | 0.88 | 0.294 |
| Expression | Very early childhood (0-2 years) | 0.03 | 0.97 | 0.173 |
| At each time period, an individual was coded as exposed if they were exposed to any type of childhood adversity. | | | | |

| Table S6. CpG-level results of loci with FDR q-value < 0.2 located in promoters of sensitive period genes, using the structured life course modeling approach (SLCMA) performed by Lussier et al. (2020). | | | | | | | |
| --- | --- | --- | --- | --- | --- | --- | --- |
| **Gene set** | **Gene name** | **CpG** | **SLCMA p-value** | **Adversity** | **Selected hypothesis** | **FDR**  **q-value^a^** | **R^2^** |
| expression | KCNK2 | cg11346522 | 1.78E-04 | Neighborhood disadvantage | Very early childhood  (1.75 years) | 0.094 | 0.022 |
| closing | MMP3 | cg16466334 | 4.03E-04 | Family instability | Early childhood  (4.75 years) | 0.148 | 0.020 |
| expression | GRIN2A | cg06922606 | 6.24E-04 | Family instability | Middle childhood  (6.75 years) | 0.148 | 0.018 |
| closing | MBP | cg26457248 | 8.38E-04 | Family instability | Early childhood  (4.75 years) | 0.148 | 0.018 |
| ^a^ FDR corrections were applied to correct for looking up results at 530 CpG sites. Lussier et al. assessed associations between time-varying exposures to the same seven types of childhood adversity as the current study and genome-wide DNA methylation (DNAm) at 440,257 CpG sites using the SLCMA, which simultaneously compares competing hypotheses for the mechanisms of exposures. | | | | | | | |

**Supplementary Figures**

Figure S1. Prevalence of exposure to childhood adversity at each developmental time period before age 7 in the analytic sample (n=785).

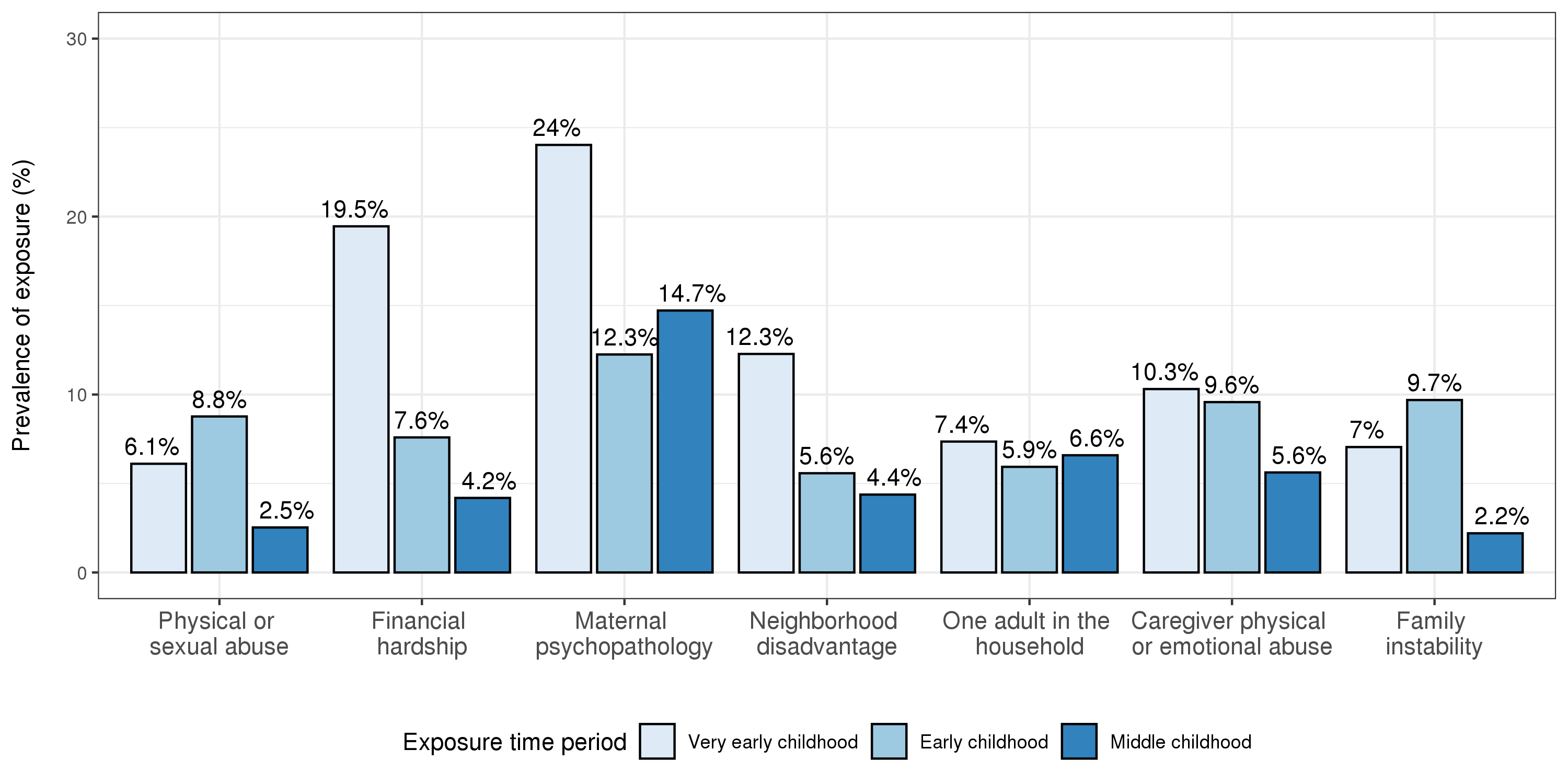

Figure S2. Tetrachoric correlations between exposures at different time periods within each type of childhood adversity before age 7 in the analytic sample (n=785). VEC: very early childhood (0-2 years); EC: early childhood (3-5 years); MC: middle childhood (6-7 years).

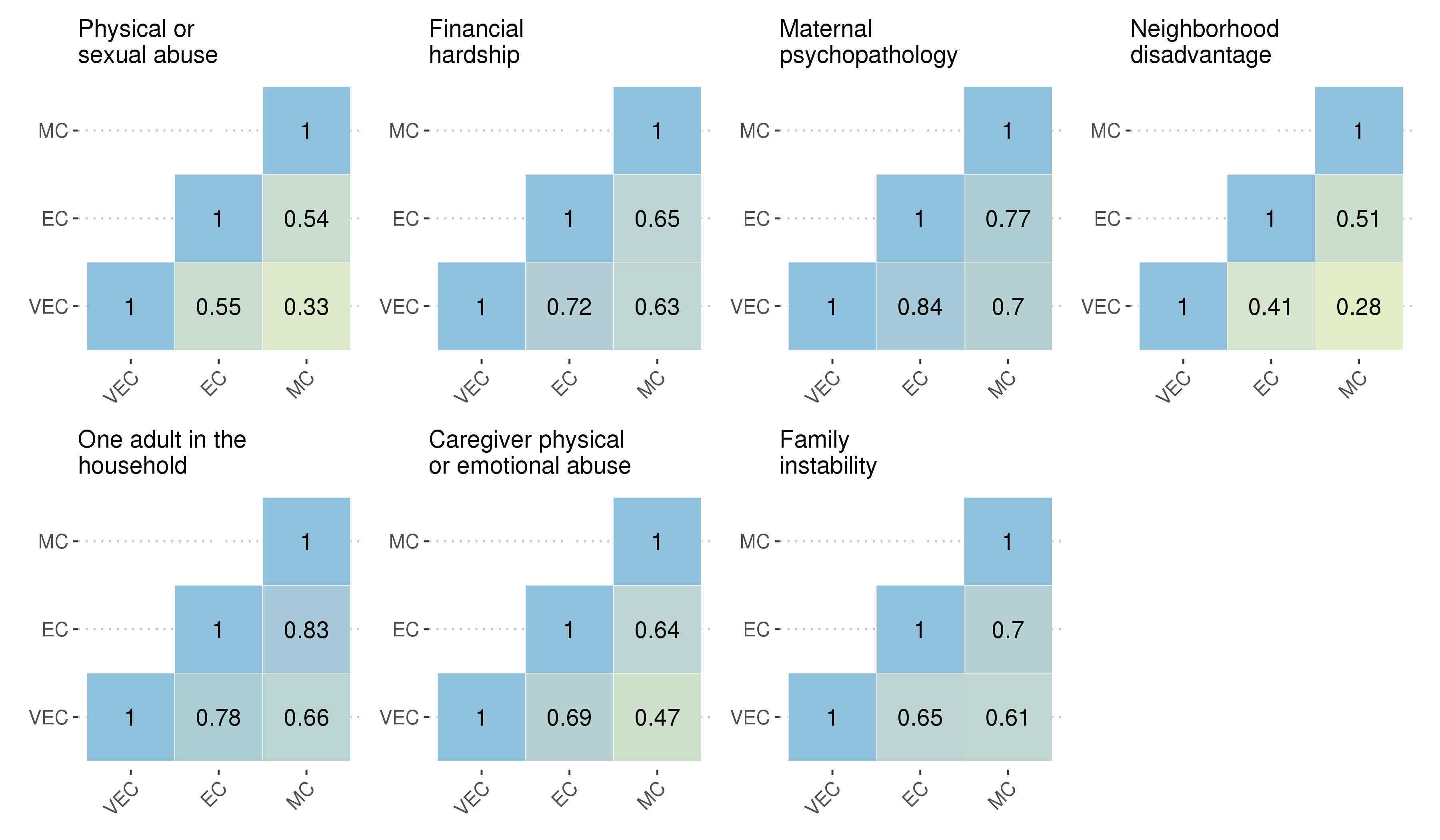

Figure S3. Tetrachoric correlations between exposures between different types of childhood adversity before age 7 within each time period in the analytic sample (n=785). VEC: very early childhood (0-2 years); EC: early childhood (3-5 years); MC: middle childhood (6-7 years).

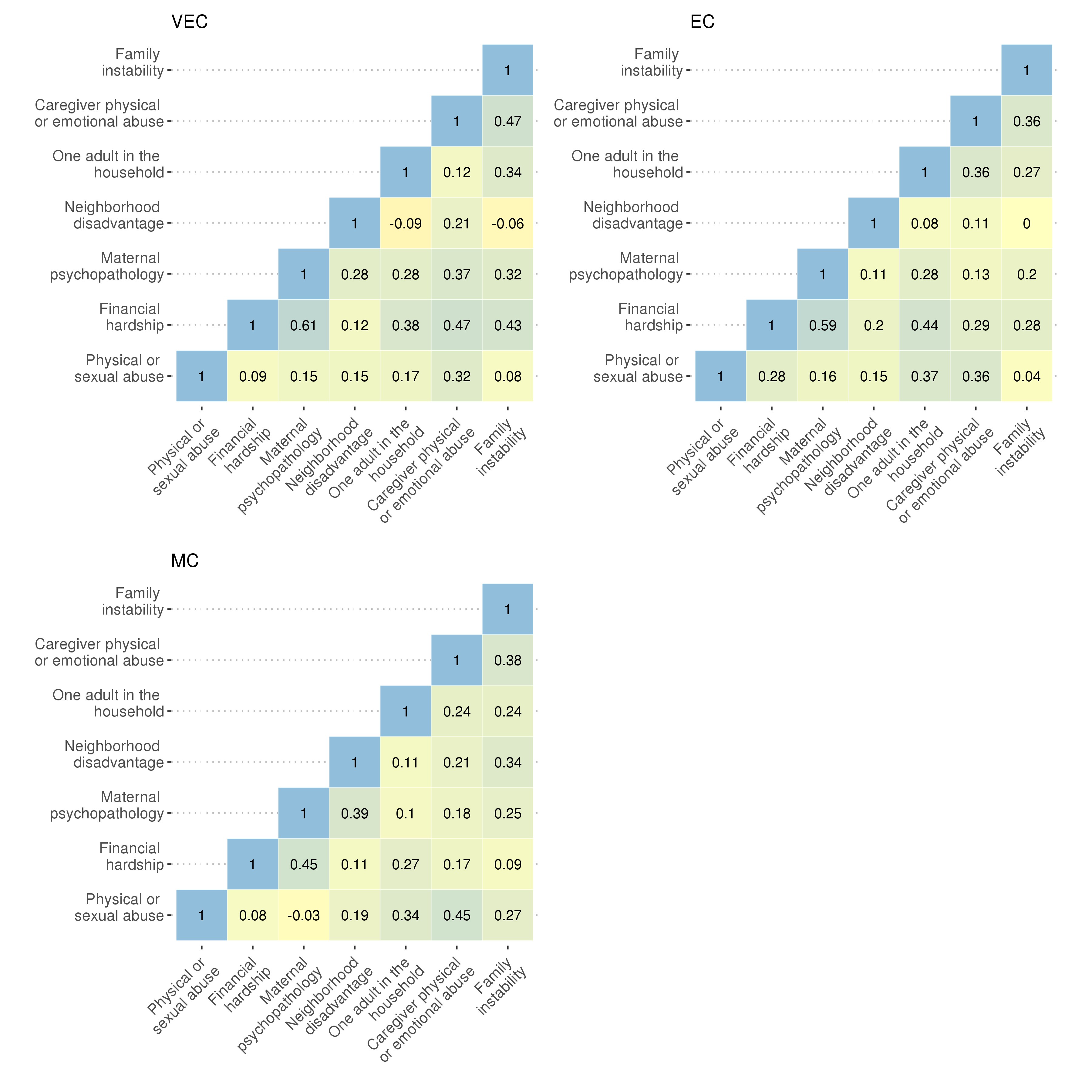
